# Technology Demonstration of a Portable Sensorimotor Assessment System after Short-Duration Spaceflight

**DOI:** 10.64898/2026.11.02.26367086

**Authors:** Rachel F. Bellisle, Elizabeth McGrath, Gordon Waddington, Lonnie G. Petersen, Dava J. Newman

## Abstract

Astronauts experience sensorimotor dysfunction due to microgravity-induced adaptations, leading to impaired posture, locomotion, and manual control. This manuscript details a portable sensorimotor assessment system, including an ankle proprioception test and visual-vestibular tests in a virtual reality headset, which was successfully deployed as a technology demonstration for one astronaut participant pre- and post-spaceflight. Post-flight testing occurred 1 and 3 days after return to Earth (R+1, R+3), with ankle proprioception also tested at R+0. Compared to pre-flight, left ankle proprioception was decreased at R+0 (two standard deviations below the mean of normative data) and returned to pre-flight levels by R+1. All visual-vestibular parameters showed decreased performance post-flight (below the 25th percentile of normative data) and little recovery by R+3. While future validation is required, these assessments appear sensitive to post-flight sensorimotor deconditioning. Furthermore, this technology demonstration highlights the logistical considerations and operational compatibility of the assessments post-flight.

## 1. Introduction

Vestibular function (the inner ear’s sense of head motion and spatial orientation), proprioception (the body’s sense of position and movement), and vision are critical inputs to the human sensorimotor system, contributing to postural control, locomotion, and other functional tasks. The reduced gravity environment experienced by astronauts alters sensory inputs (i.e., through the loss of gravitational cues), leading to adaptations in sensory processing and multisensory integration, which result in deficiencies in manual control tasks [1], reduced capacity for locomotion [2], and difficulties with postural imbalance [3]. Post-spaceflight deficiencies in postural and locomotor control are well documented, for both short- and long-duration periods of weightlessness [3–5], and result in changes in functional performance that may last for up to a week after returning to Earth [6,7]. These deficiencies in motor control have been associated with increased fall risk during locomotion [3] and have potential to impact the ability to complete critical mission tasks such as extravehicular activities and vehicle egress [6].

Additionally, spaceflight adaptations to the central processing of vestibular inputs can impact oculomotor control and gaze stability (e.g., through the vestibular ocular reflex), which may impact mission critical tasks involving visual observation or instrument displays, including rendezvous, docking, landing, and other vehicle control [8]. Although inflight exercise likely provides some protection from the loss of physiological conditioning [9,10], decrements in post-flight sensorimotor function occur even when astronauts have undertaken vigorous exercise as a countermeasure during the period of microgravity exposure [6]. NASA’s Human Research Roadmap identifies the “Risk of Altered Sensorimotor/Vestibular Function Impacting Critical Mission Tasks, Human Health, and Long-Term Health” as a significant risk for Lunar Landing and Mars missions, which requires mitigation [11].

NASA’s most recent sensorimotor standard measures, which are regularly collected from NASA and commercial astronauts, include a series of tests that are used for pre- and post-flight assessment of sensorimotor function in astronauts, including a tandem walk, sit-to-stand task, and recovery from fall task. These were selected as functional tasks that challenge sensorimotor control and simulate operationally critical tasks [4], providing information about how overall impairments could impact mission success. Additionally, these standard measures were designed to require minimal equipment, time, and physical space requirements, to ensure that testing could be completed in the field at or near the landing site, and as close to return to Earth as possible.

While appropriate for holistic performance assessments and preparation for future lunar missions, these functional tests are limited in their ability to isolate the specific impairments contributing to performance deficits, which are highly relevant to designing targeted countermeasures.

In this study, we used a portable, small-footprint, and rapidly-administered sensorimotor assessment system (Prism Neuro, Canberra, Australia) [12], which assays individual sensory domains (proprioception and visual-vestibular) (Figure 1). For somatosensory assessment, the Active Movement Extent Discrimination Assessment (AMEDA), which has been extensively utilized for proprioception assessment in existing literature [13–15], was used to determine a somatosensory acuity score for the participant’s ankle inversion. Ankle inversion proprioception is an informative measure because in normal gait the inversion/eversion movement plane has been shown previously to have greater instability, which is associated with fall risk [16]. The AMEDA device includes a tilting platform that allows ankle inversion in one foot to five possible angle positions. Following training, participants are presented with 50 random positions and asked to verbally identify the position. A performance score is provided to indicate ankle proprioception ability, and in prior literature, this score has been used to assess athlete performance and injury risk [13].

**Figure 1.**
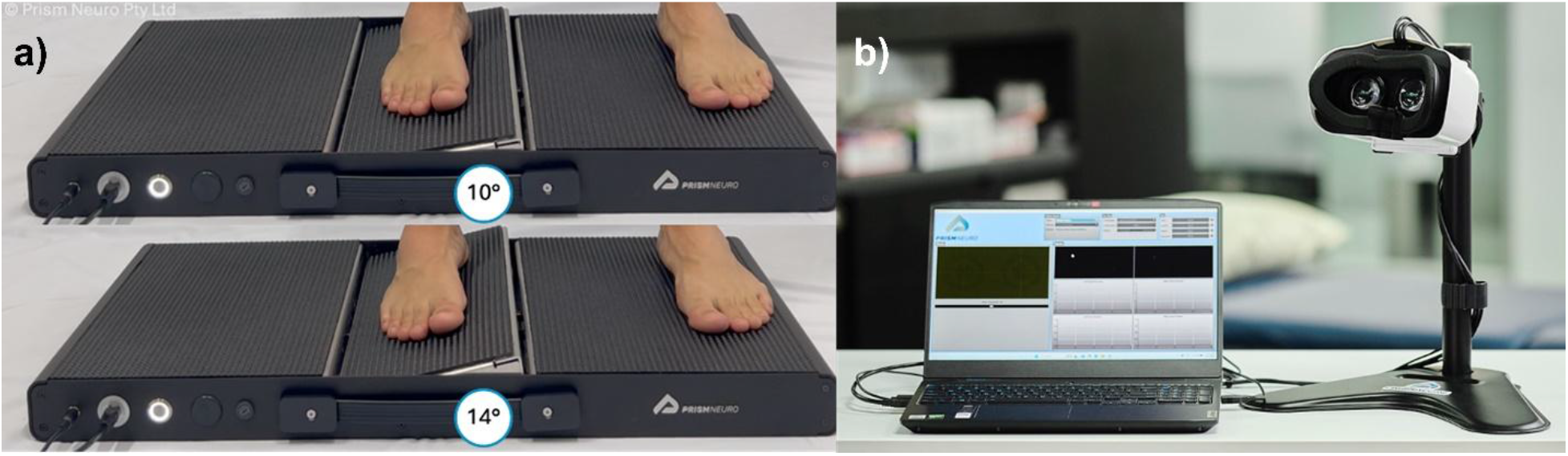
a) The AMEDA at the minimum (top) and maximum (bottom) ankle inversion positions. b) The PNEye testing equipment with tabletop virtual reality goggles.

Previous studies have shown changes in vibration sensitivity in astronauts’ feet post-flight [17,18]. Additionally, computerized dynamic posturography and sensory organization test results comparing postural sway during the disruption of different sensory inputs (e.g., eyes closed to remove vision, visual sway referencing to disrupt visual input, and support surface sway referencing to disrupt somatosensory inputs) indicate that sensory reweighting may increase reliance on somatosensory cues post-flight [19]. However, to the best of our knowledge, direct measures of ankle proprioception (e.g., AMEDA or similar approaches such as passive motion detection thresholds or joint position reproduction [20]) have never been obtained post-flight, prior to this study.

Accompanying the AMEDA assessment for proprioception, the Prism Neuro Eye (PNEye^TM^) system assesses both pupillometry and eye-tracking acuity, key functions that are indicative of sensorimotor capability [21–23]. These assessments keep the head stationary, assessing visual-vestibular integration. The PNEye system uses standard clinical evaluation methods to test the ability of a participant to accurately track a smoothly moving target using their eyes (smooth pursuit) and the ability of a participant to rapidly and accurately shift their gaze from a central to a peripheral target (saccadic targeting ability). Both of these tasks rely on the central processing capability of the vestibular complex and have been shown in the literature to identify vestibular deficits [24,25].

PNEye also incorporates the assessment of the pupillary light reflex (PLR), the changing diameter of the pupil in response to changes in light level. Features of the PLR have been shown to correlate with heart rate variability, the current gold standard for assessing autonomic function [26]. For this reason, PLR is also gaining wide acceptance as a measure of autonomic performance [27]. Autonomic function is linked to vestibular function through vestibular-autonomic responses, such as motion sickness and blood pressure regulation dependent on head position and motion, which relies on inputs from the vestibular organs [28]. Autonomic function, through control of physiologic variables, is intuitively an essential enabler of locomotor and motor control capabilities and recent neuroimaging work has demonstrated this functional reliance between motor control and physiologic control [29]. Autonomic disturbances related to altered gravity environments are also a key concern for astronaut performance [30].

This pilot study aimed to explore the operational deployment and assessment capabilities of the Prism Neuro assessment system (i.e., the AMEDA and PNEye) for spaceflight applications and leverages access to one astronaut participant before and after a 9-day mission to the International Space Station. Because this was a single-participant pilot study, the primary objective was to demonstrate the technology; the physiological findings were interpreted descriptively to inform both future validation studies and operational deployment. The work was designed to answer the following research question: Can the AMEDA and PNEye systems be successfully deployed in the field and detect changes between pre- and post-flight astronaut sensorimotor performance?

## 2. Methods

### 2.1 Participant and Study Design

One participant provided informed consent for the study protocol, approved by the NASA Johnson Space Center Institutional Review Board (Study #00000546). Demographics are not included to protect participant privacy. Data collection was completed before (L-70) and after (R+0, R+1, and R+3) a 9-day mission to the International Space Station. R+0 was completed within 3.5 hours of landing.

In-flight exercise countermeasures are not examined in this manuscript but may affect post-flight sensorimotor performance. During the mission, the participant did not have access to ISS exercise equipment but did perform exercise with simple resistance bands and used the Gravity Loading Countermeasure Skinsuit [31,32] in another portion of the study.

### 2.2 Ankle Proprioception Testing

The participant reported right leg dominance, defined as the foot used to kick a ball or write their name in the sand. The participant completed the Cumberland Ankle Instability Tool [33], a subjective test of ankle stability consisting of nine Likert-scale questions, and received a perfect score (30/30), which indicated excellent ankle stability in the participant.

Ankle proprioception was assessed during pre- and post-flight sessions, using the AMEDA (Prism Neuro, Canberra, Australia) (Figure 1a). The participant stood on the AMEDA device, which included a rotating center platform that allowed ankle inversion to five distinct angle positions (10°, 11°, 12°, 13°, and 14°) for the tested foot, while the non-tested foot rested on an adjacent, level stationary platform. During each session, the participant completed a practice session, where the study operator presented the participant with each of the 5 possible ankle positions. This was repeated 3 times before data collection began. The participant was then presented with a randomized set of 50 positions and asked to verbally indicate which position they believed they were experiencing. During the test, the participant was asked to look straight ahead, without looking at the AMEDA or their feet. The test was repeated on the left and right ankles, with a total time for practice and testing of approximately 5 minutes.

Performance on the AMEDA test was determined with an area under the curve (AUC) score derived from the receiver operating characteristic curve. Scores range from 50 to 100%, where 50% represents random chance, and 100% represents a perfect score. A previous study demonstrated that the AMEDA has moderate to good re-test reliability [12].

### 2.3 Visual-Vestibular Testing

Visual-vestibular testing occurred during pre- and post-flight sessions, using the PNEye (Prism Neuro, Canberra, Australia) testing system (Figure 1b). The system included a virtual reality (VR) headset fixed onto a tabletop with the participant seated. While initially included in the study procedure, the PNEye tests were not completed during R+0, as determined by study operators’ judgement of the participant’s post-flight condition and fatigue due to preceding tests from another study.

Upon starting the test, the participant completed a calibration process (<1 min). The VR display included a black screen with moving targets, and participants were instructed to follow a moving target with their eyes. After a successful calibration, the participant was asked to maintain their position in the VR headset for the remainder of the tests. The testing procedure (including calibration and tests) was completed in approximately 5 minutes.

Digital eye images were captured using binocular infrared video oculography at 120 fps. Gaze position and pupil diameter were recorded for each eye during each of the four tests. The tests included linear smooth pursuit, circular smooth pursuit, target-fixed PLR, and saccadic target acquisition. Six parameters were calculated: root mean square error for both linear and circular pursuit; pupil constriction latency, peak constriction velocity, and average dilation velocity for PLR; and a targeting ability ratio was calculated as average-time-on-target divided by average-time-to-target for saccadic target acquisition. A previous study demonstrated that each metric has moderate to good re-test reliability [12].

### 2.4 Statistics and Reproducibility

Due to the single-subject nature of this pilot study (*n* = 1 astronaut participant), formal hypothesis testing was not conducted. The data sets are summarized using descriptive statistics. The participant’s performance is reported using raw scores and contextualized as percentiles and z-scores relative to a previously-published normative dataset of healthy individuals (*n* = 28) [12]. This normative dataset of healthy individuals provides a terrestrial baseline necessary for contextualizing the magnitude of acute post-flight sensorimotor impairments, as well as the participant’s baseline performance.

To ensure reproducibility, PNEye calibration was performed, standardized familiarization protocols were implemented prior to data collection (e.g., three practice presentations prior to AMEDA testing), and fixed assessment volumes (e.g., 50 randomized positions per ankle) were utilized at each time point.

## 3. Results

### 3.1 Ankle Proprioception Testing

Ankle proprioception results from the AMEDA test, conducted on the left and right feet, are presented in Table 1 and Figure 2a. Pre-flight ankle proprioception performance was within one standard deviation of the normative mean for healthy individuals on Earth. Due to a pre-flight testing error (i.e., an inversion error in the participant responses, described in the discussion section), right ankle baseline data were excluded from analysis; therefore, the left ankle was used as the primary pre-flight comparator. As expected, R+0 performance declined substantially, falling to more than two standard deviations below the mean in the left ankle, and just under two standard deviations below the normative mean in the right ankle. Performance returned to or exceeded pre-flight levels by the R+1 timepoint and remained within one standard deviation of the normative mean from R+1 to R+3.

**Figure 2.**
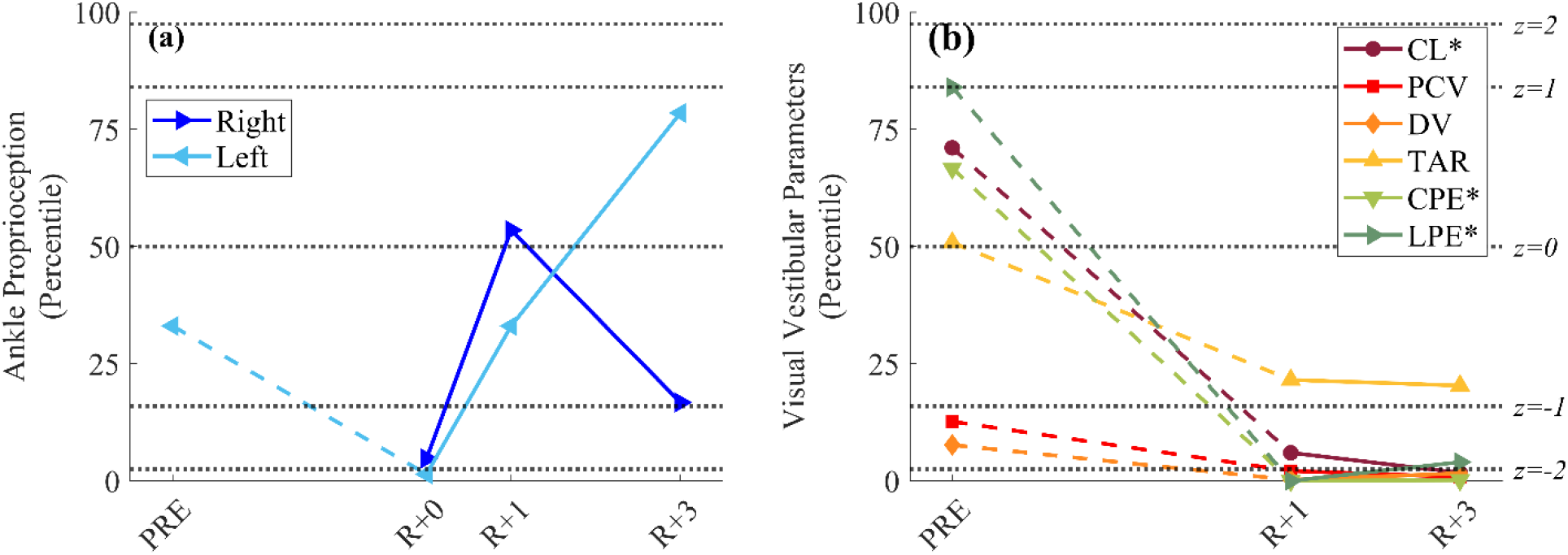
Results of a) the ankle proprioception testing and b) visual-vestibular testing at pre-flight (PRE) and post-flight time points. In the ankle proprioception testing, PRE data from the right ankle was omitted from analysis due to an inversion error in participant responses during pre-flight testing. Dotted black lines indicate the reference population mean (50^th^ percentile), and ±1 and ±2 standard deviations (z = ±1, ±2) from the mean, assuming a normal data distribution. * Some visual-vestibular parameters were inverted to allow for data visualization, where favorable performance is indicated by higher scores. CL - constriction latency; PCV - peak constriction velocity; DV - average dilation velocity; TAR - targeting ability ratio; CPE - circular pursuit error; LPE - linear pursuit error.

**Table 1.** Pre- and post-flight measures.

|  | **PRE** | **R+0** | **R+1** | **R+3** |
|---|---|---|---|---|
| **AMEDA** |  |  |  |  |
| Ankle proprioception (Left) (%) | 68 | 59 | 69 | 76 |
| Ankle proprioception (Right) (%) | - | 63 | 72 | 66 |
| **Visual-Vestibular** |  |  |  |  |
| Constriction latency, CL (ms) | 266 | - | 304 | 314 |
| Peak constriction velocity, PCV (mm/s) | 2.42 | - | 1.80 | 1.54 |
| Average dilation velocity, DV (mm/s) | 0.37 | - | 0.17 | 0.27 |
| Targeting ability ratio, TAR | 1.37 | - | 0.99 | 0.97 |
| Circular pursuit error, CPE (pixels) | 10.3 | - | 28.5 | 18.9 |
| Linear pursuit error, LPE (pixels) | 10.8 | - | 25.7 | 21.1 |

### 3.2 Visual-Vestibular Testing

Visual-vestibular parameters are presented in Table 1 and Figure 2b, along with the underlying eye tracking data for smooth pursuit testing (Figures 3a-f), PLR testing (Figures 3g-i), and saccadic target acquisition testing (Figures 3j-m). At PRE, most visual-vestibular parameters were above and within one standard deviation of the normative mean, except for peak constriction velocity and average dilation velocity, which were between one and two standard deviations below the normative mean. All visual-vestibular parameters showed a decline in performance at both R+1 and R+3 when compared with PRE (R+0 was not collected). Marginal improvements were observed in linear pursuit error, circular pursuit error, and dilation velocity at R+3 compared to R+1, but they remained under the 25^th^ percentile of the normative data and did not return to pre-flight values. There was no improvement in any other visual-vestibular parameter.

**Figure 3.**
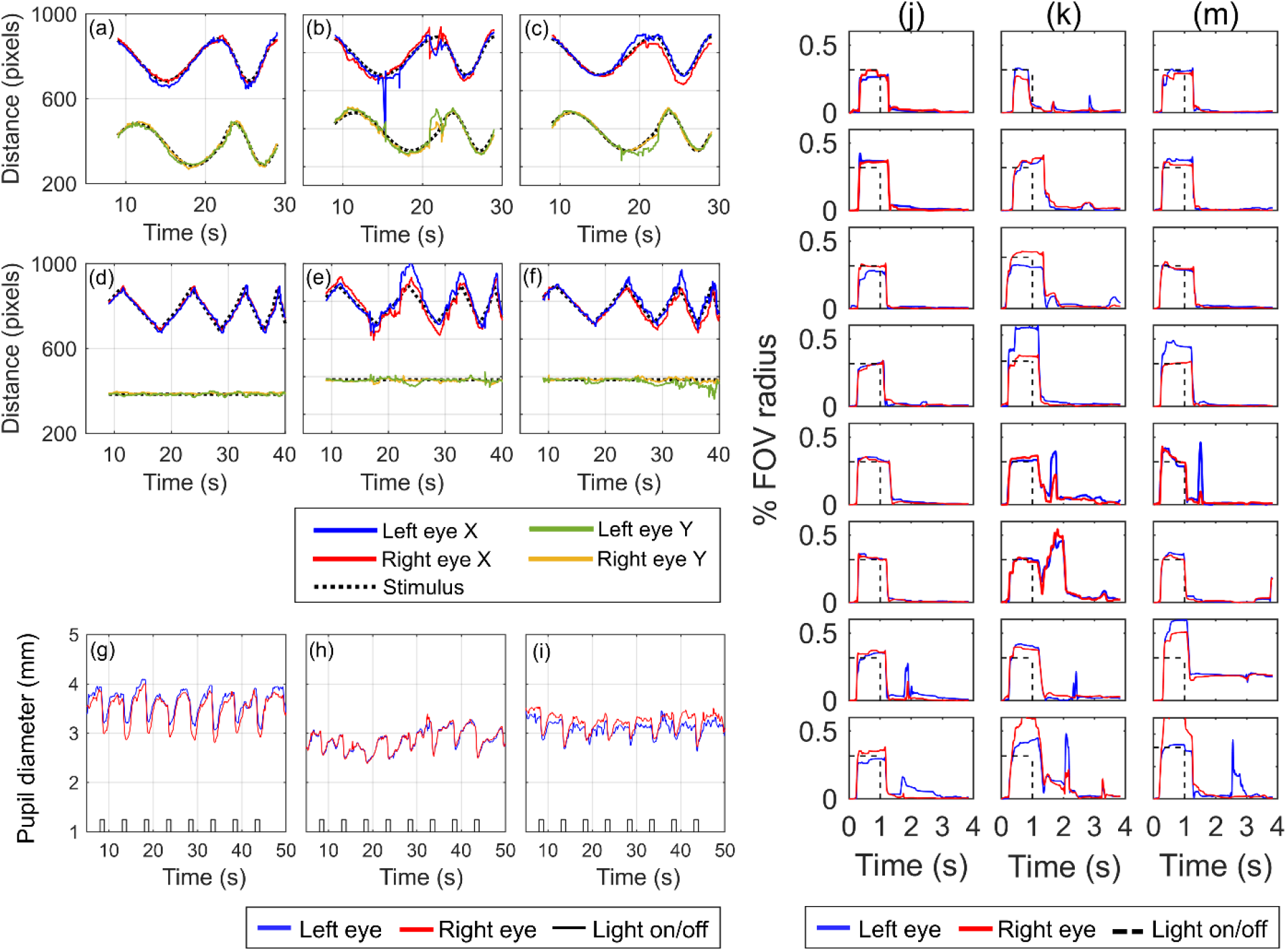
Eye tracking results. Circular smooth pursuit is shown for a) pre-flight (PRE), b) R+1, and c) R+3, and linear smooth pursuit is shown for d) PRE, e) R+1, and f) R+3. Gaze position versus time is displayed in all figures. Figures a-c show the results of the participant following a target moving in a circular pattern. Figures d-f show the results of the participant following a target moving back and forth in a horizontal line. The apexes of the sawtooth wave are the direction change points. The top traces in each pair are gaze movement in the x-direction (horizontal), and the bottom traces are the y-direction (vertical). PLR response is shown as pupil diameter versus time at g) PRE, h) R+1, and i) R+3. In each image, the participant’s gaze was focused on a central target during eight periods of light-on light-off. The resulting variation in pupil diameter is shown in the waveform. Saccadic target acquisition is also shown for j) PRE, k) R+1, and m) R+3, each for eight targets. The participant’s gaze was fixated on a central target aligned with the x-axis. A series of eight peripheral targets (dotted line) were presented to the participant for a one second interval at t = 0 s. The participant was instructed to quickly shift their focus to the target when it appeared. The waveforms show the participant’s gaze position versus time as it shifted from the central target to the peripheral target and back to the central target for each of the eight targets.

Figures 3b-c and 3e-f show the abnormalities in smooth pursuit tracking at post-flight, compared to pre-flight (Figures 3a and 3d). Eye tracking positions show an inability to maintain accurate fixation on the moving target along with the presence of saccadic intrusions in the smooth pursuit motion. Figures 3h and 3i show a reduced PLR, with lower constriction amplitude, smaller maximum and minimum pupil diameters, and less regular response patterns, at post-flight time points compared to pre-flight (Figure 3g). Figures 3k-m show poorer performance in saccadic target acquisition post-flight, with less accuracy in reaching the target, reduced synchronization between left and right eyes, and difficulty with steady fixation due to the presence of saccadic intrusions, compared to pre-flight (Figure 3j).

## 4. Discussion

This study leveraged invaluable pre- and post-flight access to one astronaut participant to provide a preliminary technology demonstration of the proposed sensorimotor assessments. While the sample size of n=1 introduces limitations and precludes broad physiological generalizations, the primary outcome is the successful demonstration of the AMEDA and PNEye systems’ ability to capture sensorimotor data in a demanding post-flight environment. The current data are used to 1) demonstrate the efficacy of the technology using physiological data (including proprioception and visual-vestibular) in a case study format with comparisons to existing literature, and 2) provide recommendations for future operations and study.

Despite a short mission duration (9 days), all measures of sensory function completed on the Prism Neuro devices show a clear reduction at the first post-flight time point compared to pre-flight. It is possible that longer mission durations would produce more severe reductions in sensorimotor performance parameters, as Miller et al. showed that the severity of post-flight functional task performance deficits is related to mission duration [6]. Additionally, our study results show that ankle proprioception performance, as measured by the AMEDA, returned to pre-flight levels by R+1, but visual-vestibular parameters did not return to baseline within 3 days.

No direct measures of ankle proprioception (i.e., AMEDA or similar approaches, such as passive motion detection thresholds or joint position reproduction [20]) have been previously obtained post-flight, prior to this study, and thus, the data reported in this paper are the first of their kind. Ankle proprioception results demonstrate the device’s sensitivity in identifying post-flight proprioceptive deficits. On the left foot, the AUC score at R+0 dropped below 62, an AUC score threshold that may indicate an increased risk of injury on Earth [34] and is approximately two standard deviations below the normative mean for healthy individuals on Earth. This score indicates that deficits in proprioception function alone may be sufficiently low to present fall risk upon return to Earth, even without considering contributions of altered vestibular function.

Previous evidence from computerized dynamic posturography testing indicates that post-flight sensory reweighting may increase reliance on somatosensory cues [19]; however, our results suggest that in the early post-flight time period, these somatosensory cues may also be severely compromised. Interestingly, a previous study showed that AMEDA results are not correlated with the somatosensory score from computerized dynamic posturography (which is commonly used post-spaceflight), indicating that these tools likely assess different aspects of somatosensory function [35].

Participant availability for pre- and post-flight testing was limited, representing a key study limitation. One AMEDA trial was completed per lower limb at each time point, with only a single pre-flight measurement session. Previous work has shown that allowing participants to perform the five required movement extents three times with limb-specific feedback provides sufficient task familiarization [36]. This familiarization method was used at the start of each testing session in our study. However, as noted in the results section, an inversion error in the participant’s responses during one test resulted in the loss of right ankle data at the pre-flight time point. Inversion errors may occur when a participant experiences a mental reversal of the AMEDA ankle position mapping. This may arise from insufficient task familiarity despite the standard practice period and is only detected during data analysis. Notably, the participant appeared to self-correct their mental model for all subsequent tests.

By R+1, AMEDA performance returned to pre-flight levels. This trend of increasing performance continued in the left foot at R+3, exceeding pre-flight levels. The unexpectedly high AMEDA score at R+3 may suggest that the pre-flight score was artificially low due to task unfamiliarity, meaning that performance at R+1 may have actually remained below the participant’s true baseline. To address these limitations, future studies should incorporate multiple pre-flight measurement sessions to improve task familiarity and reduce the risk of scale interpretation errors, which is particularly important given the high-stakes nature of astronaut assessments. Adopting a recently validated AMEDA protocol using 25 trials instead of 50 would also allow more time for familiarization while reducing overall testing fatigue [37].

The high-fidelity raw eye tracking data captured by the PNEye system also demonstrated the system’s sensitivity to post-flight reductions in smooth pursuit and saccadic targeting abilities. Similar to the ankle proprioception results, the visual-vestibular parameters showed reduced performance post-flight. Across all tested smooth pursuit parameters, the PNEye test detected a decrease in performance at R+1 compared to pre-flight performance (R+0 was not collected), with each parameter dropping below two standard deviations below the normative mean for healthy individuals on Earth. The saccadic target acquisition parameter showed a more modest post-flight decrease in performance, remaining within one standard deviation of the normative mean. These observed reductions in gaze control could have impacts on manual control and other mission critical tasks that rely on visual indicators or instrument displays [38].

Additionally, there is little recovery in smooth pursuit and saccadic target acquisition parameters by R+3. The reduced visual-vestibular performance persisted to R+3, indicating that the participant’s function had not recovered to pre-flight levels. Smooth pursuit parameters showed minor improvement at R+3 compared to the values recorded at R+1; however, these parameters remained below the 10^th^ percentile of healthy participant scores, with circular smooth pursuit error more than 2 standard deviations below the mean score of healthy participants. The saccadic targeting parameter did not improve from R+1 to R+3. These remaining deficits at R+3 are particularly striking given the participant’s short duration in microgravity.

The raw eye tracking data also indicate that both smooth pursuit and voluntary saccadic targeting abilities are compromised post-flight. During smooth pursuit the traces show a reduced ability to accurately maintain gaze on the moving target, with saccadic intrusions interrupting what should be a continuous and smooth tracking response. Similar instability is visible in the saccadic targeting traces, with reduced accuracy in target acquisition, an increase in disconjugate motion, and the presence of saccadic intrusions. These features may reflect disruption in central oculomotor processing pathways that normally support gaze fixation, saccade timing, and suppression of inappropriate saccades. In particular, the presence of saccadic intrusions is consistent with reduced inhibitory control over saccade generation, potentially involving brainstem mechanisms that regulate gaze transitions between static fixation, smooth pursuit, and voluntary saccades [39].

Our PNEye results reveal persistent oculomotor deconditioning, with some differences from existing literature. Previous qualitative oculomotor tests of 14 astronauts within 2 hours of landing from week-long Space Shuttle missions detected no clinical abnormalities in saccade or pursuit tracking [40]; however, our quantitative assessment detected impairments that persisted several days post-flight. It is possible that the quantitative PNEye system provides a more sensitive measurement than qualitative observation. Quantitative electro-oculography (EOG) during Space Shuttle missions showed inflight oculomotor changes, including increases in horizontal saccadic latency and reduced peak velocities. Saccade velocity, the only metric available post-flight, had largely returned to pre-flight values within ∼1 h of landing [41]. The comparison between the previous study and ours is limited as they use different measurement tools and quantify different aspects of saccadic performance; however, our results show a differing trend for reduced targeting ability ratio persisting to R+1 and R+3.

Similar to the PNEye approach, several previous studies have used eye tracking to measure oculomotor task performance. In-flight measures on cosmonauts during long-duration missions (76-438 days) showed an increase in the number of corrective saccades during a target tracking task, and during smooth pursuit tasks, eye movements were stepwise saccadic motions rather than smooth tracking [42]. Similarly, the post-flight PNEye smooth pursuit tasks showed some corrective saccades and greater tracking errors. Another study of Russian cosmonauts during visual-manual tracking tasks showed post-flight decreases in visual tracking accuracy, with recovery to pre-flight levels by 8–9 days after landing [43], showing a recovery timeline extending beyond our R+3 timepoint.

Comparisons with previous studies also indicate a distinction between head-fixed and head-eye coordination tasks. Studies of head-free oculomotor tasks, which use integrated eye-head coordination strategies, have demonstrated significant post-flight decreases in smooth pursuit tracking gain and reduced head and eye velocities [44], and increased gaze latencies during target acquisition [45]. However, Reschke, et al. showed recovery of target acquisition deficiencies at R+2, while our work does not show any meaningful recovery by R+3. The lack of recovery observed in our isolated gaze control tasks (where the head was kept stationary) may suggest distinct deconditioning and recovery profiles for oculomotor control during gaze control when isolated from active head-movement compensation, which may be an appropriate topic for future study with the PNEye system.

Smooth pursuit and saccade tasks are often used to assess traumatic brain injury and concussion, and the resulting disruption to central processing may have some similarities to the results shown in this study. Lunkova et al. showed, using fMRI, that smooth pursuit and saccade oculomotor tasks produced brain neural activation patterns that aligned with regions of the brain affected by concussion [46]. Like the participant in this study, oculomotor results in acute concussion patients exhibit deficits in smooth pursuit tasks and saccade tasks [47], and oculomotor measures are highly predictive of acute concussion [48].

In addition to visual tracking tasks, PLR parameters decreased at R+1 compared to pre-flight performance (R+0 was not collected). PLR is often used as a measure of autonomic function [26], and the lack of recovery for these metrics may indicate ongoing autonomic deficits post-flight. Peak constriction velocity and average dilation velocity dropped below two standard deviations below the normative mean for healthy individuals on Earth. Constriction latency dropped between one to two standard deviations below the normative mean. While constriction latency was above and within one standard deviation of the mean at the pre-flight timepoint, peak constriction velocity and average dilation velocity were already between one to two standard deviations below the normative mean, indicating that even pre-flight the participant may have experienced mild autonomic dysfunction. However, the subsequent post-flight decline indicates a further decrement in autonomic responsiveness after spaceflight.

Two of three PLR parameters did not show any improvements from R+1 to R+3. While some marginal improvements were seen in dilation velocity between R+1 and R+3, it remained more than two standard deviations below the mean score of healthy participants. Overall, the reduced autonomic performance persisted to R+3, and the participant’s function did not recover to pre-flight levels by our last data collection session.

Our post-flight PLR findings are consistent with the only other quantitative pupillometry dataset from human spaceflight of which we are aware. Using a handheld automated pupillometer in four astronauts on a short-duration mission, Shirah et al. reported that constriction and dilation velocities rose above pre-flight baseline in microgravity and then fell below baseline on return to 1G [49]. Our results, obtained with a different instrument, show the same post-flight direction of change. That work attributed these changes primarily to headward fluid shifts and intracranial pressure rather than to autonomic function; the present data cannot distinguish between these mechanisms.

Our findings are also consistent with established effects of spaceflight on autonomic function. Post-flight orthostatic intolerance is commonly observed in astronauts and likely results from microgravity-induced changes in fluid volume and the adaptation of the autonomic nervous system [30], including altered vestibular cues which provide critical signals for the autonomic function of blood distribution [50,51]. Prior work has also indicated that an increased heart rate during an orthostatically challenging rapid standing task is correlated with decreased postural stability, indicating the connection between balance control and cardiovascular responses [52]. Post-flight orthostatic intolerance timelines show that heart rate (measured after a passive position change to upright) did not recover within 10 days post-flight in 12 astronauts who completed 6-month missions [50], which is consistent with our study’s lack of PLR recovery within 3 days of return to Earth. Other research for military aviators has shown that training to reduce motion sickness attenuated the decline in PLR parameters following vestibular provocation, supporting the potential value of PLR as an objective marker of autonomic adaptation in aerospace settings [53].

Beyond the physiological sensitivity results, the study also provided practical insight into the feasibility of deploying these assessments operationally. Operators found that the small footprint, quick set-up time, and easy transport were advantageous for R+0 testing at the field site. These qualities suggest that the AMEDA and the PNEye are compatible with the logistical needs of post-flight testing. The PNEye system has also been designed to be well-tolerated, using a seated position with the head stationary to alleviate potential nauseogenic effects associated with VR headsets. However, R+0 verification is still needed. While the equipment was present at R+0, the PNEye test was not conducted at R+0 due to concerns for participant comfort and the high post-flight testing load, which resulted in fatigue from preceding tasks from another study. The decision to omit this test was made at the discretion of the operator and underscores a critical operational constraint: the necessity of carefully balancing testing volume against the astronaut’s exceptionally high physical and cognitive load during post-flight recovery. Future testing should ensure that testing volume and scheduling is sufficient for all included measures. As discussed previously, scheduling multiple pre-flight data collection sessions (for both the AMEDA and PNEye tests) is also recommended for future use, to ensure task familiarity and the availability of reliable pre-flight data.

While current sensorimotor standard measures are completed pre- and post-flight, there are currently no inflight assessment tools used to monitor sensorimotor performance between launch and return to Earth, apart from tools still in development [54,55]. Unlike current sensorimotor standard measures, the AMEDA and PNEye tests potentially do not require upright posture, and thus, operations in micro- and partial-gravity are likely feasible. However, additional testing is required to understand 1) the impact of gravity environment on the testing parameters and their relation to upright performance on Earth [54], and 2) in-flight ISS operational feasibility as considered in other proposed sensorimotor assessment tools for spaceflight applications [56]. Some validation has already been completed for the AMEDA during acute body unloading to simulate micro- and partial gravity [14,57], and similar tools show how this type of device may be used in a microgravity environment with vertical weight loading [54,55]. As VR systems are already used during space missions aboard the ISS, the visual–vestibular testing components may be adaptable for delivery through existing onboard systems, potentially reducing the need for additional equipment. The PLR component, while used in the present context as an indicator of autonomic function, has also been investigated as a potential marker relevant to intracranial pressure (ICP), which is implicated in spaceflight-associated neuro-ocular syndrome (SANS) [58]. PLR testing may therefore provide multifunctional outputs, increasing the efficiency of routine crew monitoring. Automated pupillometry has already been deployed aboard the ISS during a short-duration commercial mission, where a handheld device was used to collect pre-flight, in-flight, and post-flight measurements from four crewmembers, further supporting the operational feasibility of automated PLR assessment in spaceflight [49].

This study provides a preliminary demonstration of the AMEDA and PNEye sensorimotor assessment systems in an astronaut pre- and post-flight, and demonstrated that these measures are sensitive to post-spaceflight sensorimotor deficits. Further validation of this sensorimotor assessment system with a larger participant cohort is necessary, but future work may also investigate whether optimizing the sensorimotor and autonomic metrics with targeted interventions as part of pre-mission or in-mission training would provide additional resilience to the in-mission effects of low gravity. As humans return to the moon for the first time in over 50 years, the improved understanding of sensorimotor function after exposure to microgravity is critical to ensuring safety and reducing injury risk during critical mission tasks on the lunar surface and informing relevant countermeasures. Overall, the results of this work encourage further investigation of the proposed sensorimotor assessment system in astronaut populations and aim to support assessment capabilities to improve the understanding of the effects of spaceflight on the human body in preparation for partial gravity operations and safe return to Earth.

## Data Availability

The AMEDA datasets generated and analyzed in this study are available on reasonable request. Raw data traces from visual-vestibular tasks are included in this manuscript.

## Declaration of generative AI and AI-assisted technologies in the manuscript preparation process

During the preparation of this manuscript, the authors utilized Gemini 3.1 Pro and Claude Opus 5 to assist with editing and proofreading. Following the use of these tools, all authors thoroughly reviewed, edited, and validated the manuscript’s text and take full responsibility for the final content and scientific accuracy of the publication.

## Acknowledgments

RB was supported by a Draper Scholarship with The Charles Stark Draper Laboratory, Inc. The authors thank the MIT Space Exploration Initiative (Sean Auffinger and Ariel Ekblaw) and the MIT Apollo Program Professor of Astronautics Chair for funding and mission integration support. We thank the private spaceflight partners for supporting this experiment. Thank you to Golda Nguyen for data collection support, and thank you to our study participant.

## Author Contributions

RB: Conceptualization, Methodology, Investigation, Formal analysis, Writing – original draft, Writing – review & editing. EM: Formal analysis, Methodology, Writing – review & editing. GW: Methodology, Writing – review & editing. LP: Methodology, Writing – review & editing. DN: Conceptualization, Supervision, Writing – review & editing.

